# Separate Fever Clinics Prevent the Spread of COVID-19 and Offload Emergency Resources: Analysis from a large tertiary hospital in China

**DOI:** 10.1101/2020.04.03.20051813

**Authors:** Jiangshan Wang, Liang Zong, Jinghong Zhang, Han Sun, Joseph Harold Walline, Pengxia Sun, Shengyong Xu, Yan Li, Chunting Wang, Jihai Liu, Fan Li, Jun Xu, Yi Li, Xuezhong Yu, Huadong Zhu

**Author notes:** **Corresponding author:** Huadong Zhu, **Address for correspondence:** Department of Emergency Medicine, Peking Union Medical College Hospital, No.1 Shuaifuyuan Wangfujing, Dongcheng District, Beijing, China, **Email:**, **Co-Corresponding author:** Jihai Liu, **Address for correspondence:** Department of Emergency Medicine, Peking Union Medical College Hospital, No.1 Shuaifuyuan Wangfujing, Dongcheng District, Beijing, China, **Email:**. These authors contributed equally to this work (Jiangshan Wang, Liang Zong).

## Abstract

**Objectives:** COVID-19 began spreading widely in China in January 2020. Outpatient “Fever Clinics” (FCs), instituted during the SARS epidemic in 2003 were upgraded to provide COVID-19 screening and prevention attached to large tertiary hospitals. We sought to analyze the effect of upgraded FCs to detecting COVID-19 at our institution.

**Design:** A population-based cross-sectional study.

**Participants:** A total of 6,365 patients were screened in the FC.

**Methods:** The FC of Peking Union Medical College Hospital (PUMCH) was upgraded on January 20, 2020. We performed a retrospective study of patients presenting to the FC between December 12, 2019 to February 29, 2020, covering a period of 40 days before and after upgrading the FC. All necessary data, including baseline patient information, diagnoses, follow-up conditions for critical patients, transfer information between the FC and emergency department (ED) were collected and analyzed.

**Results:** 6,365 patients were screened in the FC, among whom 2,192 patients were screened before January 21, 2020, while 3,453 were screened afterwards. Screening results showed that upper respiratory infection was the major disease associated with fever. Compared to before the outbreak, patients transferred from the FC to ED decreased significantly [39.21% vs 15.75%, p<0.001] and tended to spend more time in the FC [55 vs 203mins, p<0.001]. For critically-ill patients waiting for a screening result, the total length of stay in the FC was 22mins before the outbreak, compared to 442mins after the outbreak (p< 0.001). The number of in-hospital deaths of critical-care patients seen first in the FC was 9 of 29 patients before the outbreak and 21 of 38 after (p<0.050). Nineteen COVID-19 cases were confirmed in the FC, but no other patients or medical care providers were cross-infected.

**Conclusion:** The work-load of the FC increased after the COVID-19 outbreak and effectively prevented COVID-19 from spreading in the hospital,as well as offload ED resources.

## Introduction

### Background

Coronavirus disease 2019 (COVID-19) caused by the severe acute respiratory syndrome coronavirus 2 (SARS-COV-2) broke out in Wuhan, Hubei Province at the end of 2019^[1]^, and cases are now rapidly spreading worldwide ^[2]^. Currently, controlling the spread of SARS-CoV-2 is of primary concern ^[3]^. The main manifestations of this disease include acute fever, cough and dyspnea ^[4]^, thus emergency departments (EDs) have become the primary facilities providing initial diagnoses and medical care for potential COVID-19 patients. Unfortunately, as the virus spreads widely, crowded patients in EDs face a high risk of cross-infection ^[5,6]^. In mainland China, outpatient “fever clinics” (FCs), affiliated to the ED, are designed to help separate potentially infectious from non-infectious patients ^[7]^. FCs were started at the suggestion of National Health Commission of the People’s Republic of China as early as 2003 during the SARS outbreak in China Even after the SARS event, FCs were still preserved as a location near EDs for early identification and isolation of potentially infectious patients ^[8]^. Therefore, few suspected patients were managed in emergency department^[9]^. However, between SARS in 2003 and the current COVID-19 outbreak, the FC system has seen few similar stresses and few reports have emerged about this potentially key element of hospital infection-prevention infrastructure.

### FC upgrade after the COVID-19 outbreak

Before the COVID-19 outbreak, four doctors were allocated to the FC where influenza A and B were screened for every patient suffering from both fever and respiratory symptoms. The FC was also tasked with excluding eruptive infectious diseases (e.g. measles, rubella, and varicella). Patients with infectious diseases received initial therapy in the FC, and then were transferred to inpatient isolation wards if needed; other patients were transferred to the ED. After the COVID-19 outbreak, as many as twelve doctors wearing “grade-3” isolation gowns worked in the FC^[10]^. Two consulting rooms were added to supplement the original single-room. The number of medical care providers providing in-person coverage every 24 hours increased from two to nine every 24 hours, while nursing staff increased from nine to 15 every 24 hours. Rescue equipment such as endotracheal intubation tools, central venous catheters, noninvasive and invasive ventilator machines, high-flow oxygen therapy devices and bedside ultrasound were added or expanded.

All patients with either fever or respiratory symptoms, no matter with or without a history of Covid-19 exposure, were mandated to go through FC triage (see Figure 1). Each patient was required to wear a mask on arrival to the FC and was allocated to different regions according to their triage history and clinical severity (see Figure 1). FC took responsibility for screening SARS-COV-2, in addition to influenza and eruptive diseases noted above. All acquired nucleic acid samples were tested by two independent laboratories that had been authorized by the Beijing Municipal Health Commission. Only “double negative” results was defined as a negative result for the patient. In addition to identification, there were specialized doctors in charge of suspected patients, critical patients and common patients, respectively. Negative pressure isolation wards with complete sets of resuscitation equipment were readied for any critical patients. Once the screening tests were reported, confirmed patients would be transferred to specialized hospitals whereas others who needed further treatment were transferred into the ED (see Figure 2).

**Figure 1.**
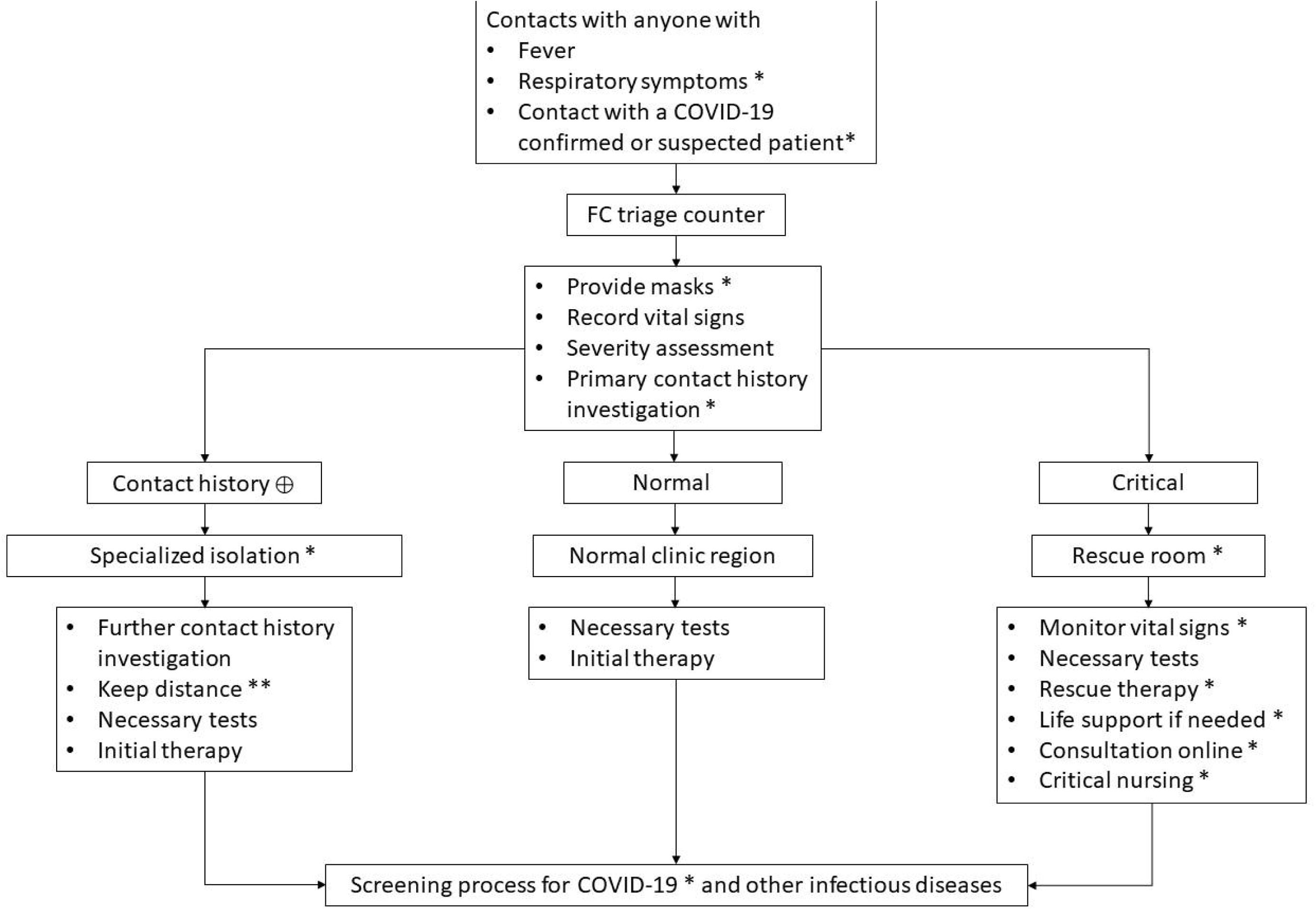
Triage process and regional isolation for different patients in the FC. * Upgraded FC parts. ** Educate patients to maintain a person-to-person distance greater than two meters. Abbreviations: FC: fever clinic, ⊕: positive, COVID-19: coronavirus disease 2019.

**Figure 2.**
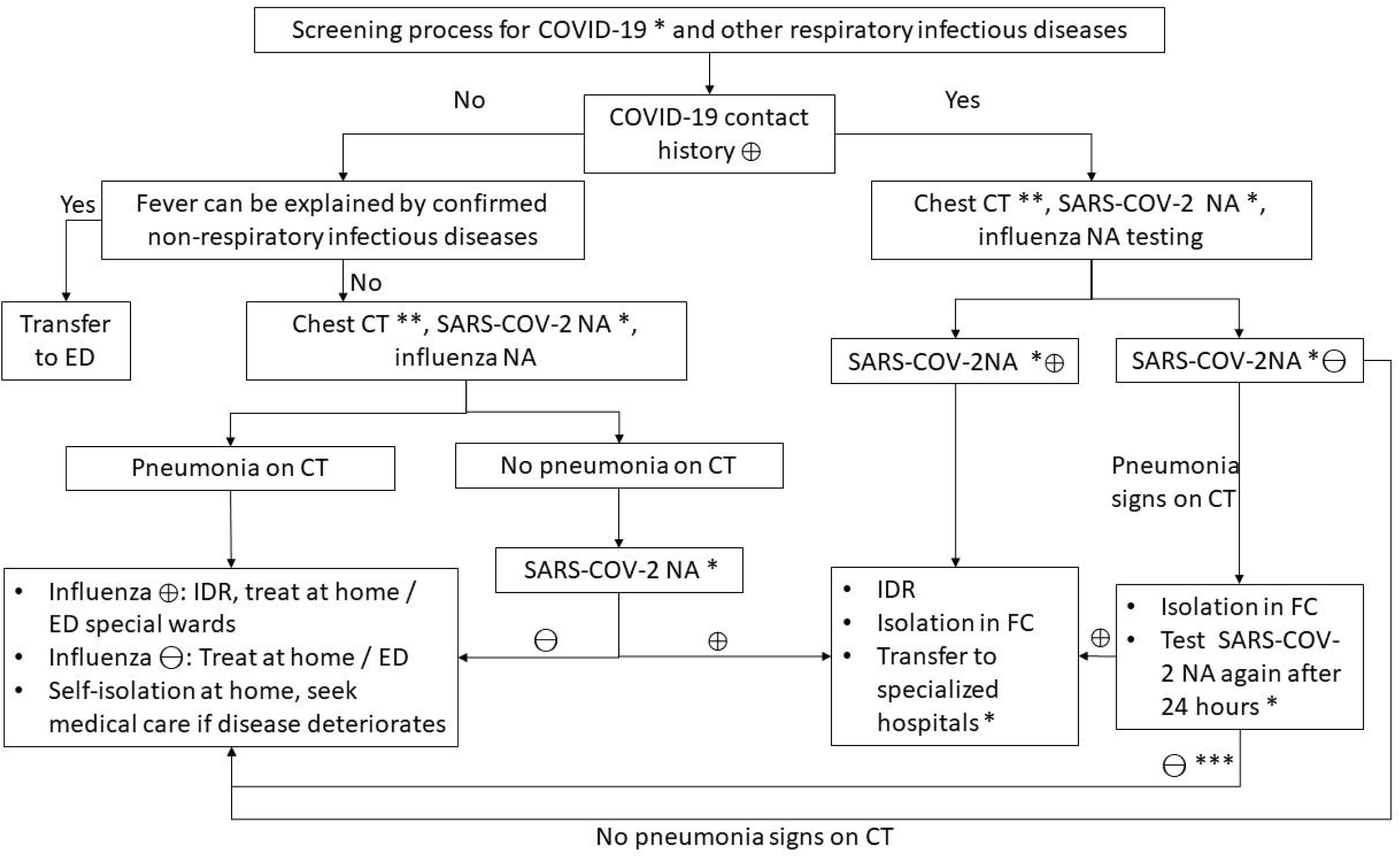
Screening process for COVID-19 and other respiratory infectious diseases. * Upgraded FC parts. ** After the COVID-19 outbreak, Chest CT was suggested as a routine examination for patients in FC excluding special populations such as pregnant women and children. *** Patients are recommended to test for SARS-COV-2 RNA again one week later even if previous RNA tests were negative twice if they have: (1) confirmed COVID-19 contact history; (2) clinical manifestations and lab tests implying viral infection which can not be explained by other diseases; (3) chest CT strongly suggestive of viral pneumonia. Abbreviations:, COVID-19: coronavirus disease 2019, ⊕: positive, ⊖: negative, CT: computed tomography, SARS-COV-2: severe acute respiratory syndrome coronavirus 2, NA: nucleic acid, ED: emergency department, IDR: infectious disease report, FC: fever clinic.

## Methods

### Data collection

We collected data from all patients who presented to the FC of PUMCH 40 days before the upgrade in the FC (December 12, 2019 to January 20, 2020), and for 40 days after the upgrade from January 21 to February 29, 2020. The FC upgrade date (January 20, 2020) was also the official date Covid-19 was declared an “outbreak” in Beijing. We included all critically ill patients during this period who presented to the FC and then were transferred to the ED. The data were collected from patients’ medical records and their registration information at the time of presentation. Patients’ clinical condition, primary diagnoses, time of registration (FC and, potentially, ED), as well as the duration of consultation at each visit were obtained.

Critically ill patients were identified according to the following criteria: (1) patients transferred to resuscitation rooms in the ED from the FC after initial screening and initial treatment; (2) APACHE II score ≥8; (3) patients who were ruled out the possibility of COVID-19 pneumonia^[11]^. Critically ill patients’ prognoses and treatment results were documented by medical records, as well as any changes in patients’ condition within seven days following initial presentation to the FC (improvement, non-improvement or death).

#### Patient and Public Involvement

This was a restrospective study,we collected medical information of all involved patients from electronic information system.The patients did not involve in the recruitment to and conduct of the study, as well as not join in designing.

### Statistics

Statistical Package for Social Sciences 24.0 software (IBM Corp., Aramonk, NY, USA) was used for statistical analysis. The Kolmogorov-Smirnov test was used to determine the normal distribution of variables. Variables with normal distribution are shown as a mean (±SD). T-tests were used for variables that followed normal distribution.. Data that did not follow normal distribution was shown as a median (25%-75%) and analyzed by the Wilcoxon rank sum test. Chi-square tests and Fisher’s exact tests were used for enumeration data. P-values less than 0.05 were taken to indicate statistical significance.

## Results

### Patient Characteristics and Disease Etiologies

In total, 6,365 patients were screened in the FC, among whom 2,192 patients were screened before the outbreak, and 3,453 patients were screened after the outbreak was declared and the FC upgraded on January 20, 2020. There was no statistical difference in sex ratio between the two groups, but a significant difference in age was seen (p=0.001). The most common disease found in the FC was an upper respiratory infection, followed by an abdomino-pelvic infection and pneumonia (see Table 1).

**Table 1:**
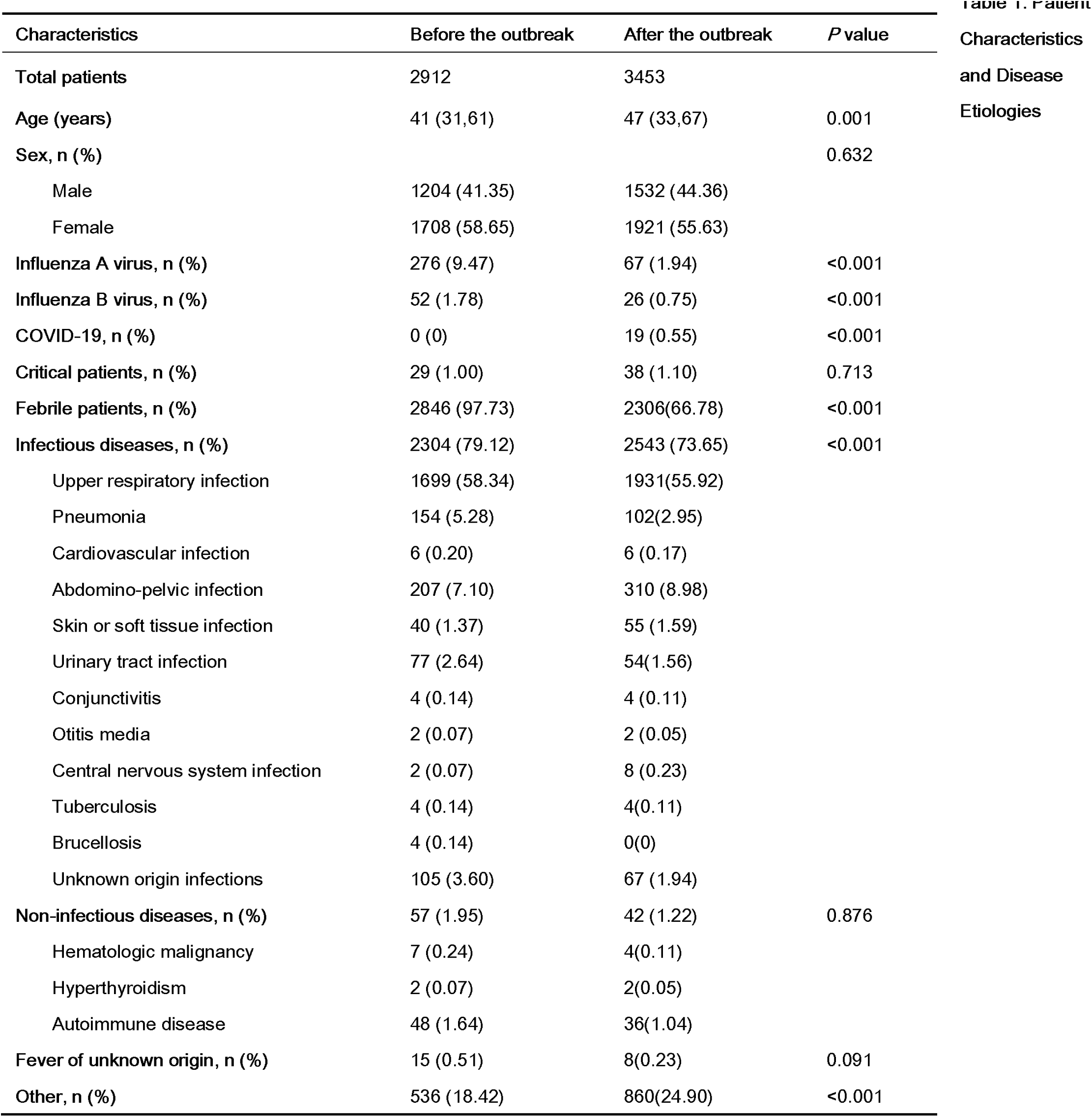
Patient Characteristics and Disease Etiologies

### FC to ED transfer statistics before and after the COVID-19 outbreak

The number of registered patients who presented first to the FC and were then transferred to the ED before and after the outbreak was 1,142 and 544, respectively. There was no statistical difference in the sex ratio or age of patients between these two groups (p>0.05). 1,083 (94.84%) of cases before the outbreak took less than 24 hours to transfer between the FC and ED. After the outbreak, 482 cases met this benchmark (p<0.001). Meanwhile, the treatment time in the FC grew significantly longer compared to before the outbreak (p<0.001) (see Table 2).

**Table 2:**
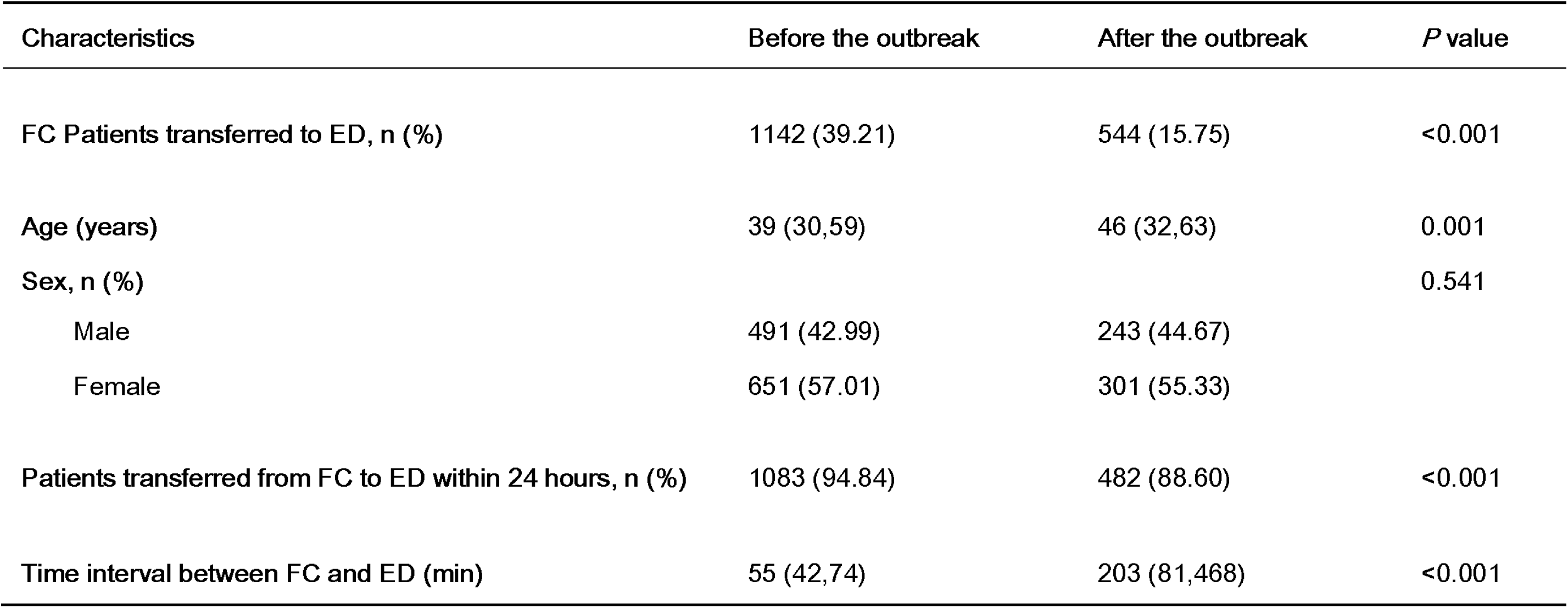
FC to ED transfer data before and after the COVID-19 outbreak

### Critically ill patients

69 critically ill patients were included in our analysis, and two patients were excluded due to an APACHE II score < 8 points. 29 and 38 patients presented, respectively, at the FC before and after the COVID-19 outbreak. The ratio of male to female patients was 1.23:1, average age was 61 (44,76). There was no significant difference in the sex ratio or age of patients between the two groups (p>0.05). There was also no significant difference in the severity of disease between the two groups when examining the respective APACHE II scores (16.1±6.67 vs 18.74±6.72 (p>0.05). Patients with septic shock and pneumonia combined with respiratory failure accounted for most diagnoses. The number of in-hospital deaths within seven days for critically ill patients initially presenting to the FC was 9 of 29 and 21 of 38 before and after the outbreak, respectively (p<0.05) (see Table 3).

**Table 3.** Characteristics and Disease Etiologies of Critically Ill Patients

| Characteristics | Before the outbreak | After the outbreak | *P* value |
| --- | --- | --- | --- |
| Critical patients, n (%) | 29(1.00) | 38(1.10) | 0.713 |
| Age (years) | 59.45±19.86 | 63 (47,78) | 0.735 |
| Sex, n (%) |  |  | 0.630 |
| Male | 15 (51.72) | 22 (57.89) |  |
| Female | 14 (48.28) | 16 (42.11) |  |
| APACHE II score | 16.1±6.67 | 18.74±6.72 | 0.116 |
| Diagnosis, n (%) |  |  |  |
| Fever | 29 (100) | 38 (100) |  |
| Septic shock | 9(31.03) | 12(31.58) |  |
| Pneumonia with respiratory failure | 9(31.03) | 10(26.31) |  |
| Acute myocardial infarction | 2(6.70) | 2(5.26) |  |
| Gastrointestinal bleeding | 2(6.90) | 1(2.63) |  |
| Acute cerebrovascular disease | 1(3.45) | 4(10.52) |  |
| Intracranial metastasis of lymphoma | 0(0) | 1(2.63) |  |
| Central nervous system infection | 2(6.90) | 1(2.63) |  |
| Acute myocarditis | 1(3.45) | 1(2.63) |  |
| Tachyarrhythmia | 2(6.90) | 0(0) |  |
| Ruptured iliac aneurysm | 1(3.45) | 0(0) |  |
| Acute aortic dissection | 0(0) | 1 (2.63) |  |
| Diffuse alveolar hemorrhage with SLE | 0(0) | 1(2.63) |  |
| Acute left heart failure | 0(0) | 2(5.26) |  |
| Acute pulmonary embolism | 0(0) | 1(2.63) |  |
| Hyperosmolar hyperglycemic state | 0(0) | 1(2.63) |  |
| Prognosis, n (%) |  |  | 0.021 |
| Improvement | 16 (55.17) | 17 (44.74) |  |
| Non-improvement | 4 (13.79) | 0(0) |  |
| Death | 9 (31.03) | 21 (55.26) |  |

SLE=Systemic Lupus Erythematosus

### Length of stay in FC before and after the COVID-19 outbreak

The total length of stay in the FC was 22 (12,47) mins before the outbreak, compared with 442 (374,636) mins after the outbreak (p<0.001). While the total length of stay in the resuscitation rooms of the ED lengthened from 22 (7,59) hours to 48 (21,96) hours after the outbreak (p<0.001) (see Table 4).

**Table 4:** Length of Stay in the FC Before and After the COVID-19 outbreak

| Characteristics | Before the outbreak | After the outbreak | *P* value |
| --- | --- | --- | --- |
| FC total length of stay (minutes) | 22 (12,47) | 442 (374,636) | <0.001 |
| ED resuscitation room total length of stay (hours) | 22 (7,59) | 48 (21,96) | <0.001 |
| Treatment times (minutes) | 165(95,241) | 123 ( 78,164 ) | 0.072 |
| Supportive treatment times (minutes) | 153.2±100.3 | 72 (34,160) | 0.065 |

Most commonly provided treatments in the FC were: antibiotics, antiarrhythmic drugs, antihypertensive drugs, and antiplatelet drugs. Common supportive treatments included: nasal catheter oxygen, non-invasive/invasive ventilation, fluid resuscitation, vasopressors, intracranial pressure reduction, and diuretic drugs. Initial treatment times are shown in Table 4.

### Strengths and limitations

This study had several limitations. It was a restrospective study at a single center and carries the associated weaknesses of such study methodologies. Still, given the seriousness of the global fight against COVID-19, the lessons learned through the expansion of the FC and its relationship to the ED and ED patient flow are important concerns for global discussion. Future studies should examine the effects of having dedicated FC associated with ED, including the degree of integration with the ED vs. the rest of the hospital.

## Discussion

COVID-19 is a deadly new respiratory infectious disease. This widely spreading disease became a notifiable infectious disease starting on January 20, 2020 per the National Health Commission of the People’s Republic of China. Starting on January 20, the Beijing Municipal Government initiated a level-1 (highest) public health response to prevent the spread of the disease^[12]^. PUMCH upgraded the FC that same day to enhance the screening and treatment of potential COVID-19 patients. In this study, we reviewed the details of the FC in the 40 days before and after the COVID-19 outbreak. We found that after the outbreak, more patients received treatment in FC, critically ill patients received initial rescue management in the FC, and, most importantly, no confirmed COVID-19 patients were transferred to the ED and no other patients, doctors or nurses were infected in the hospital. This FC upgrate strategy seemed to successfully prevent COVID-19 from spreading.

According to our data, upper respiratory infections were the major disease seen in the FC both before and after the COVID-19 outbreak. Most mild COVID-19 patients had upper respiratory infection syndromes ^[13]^, but they also are strongly infectious, which causes dramatic difficulties in screening. Consequently, it was not possible to exclude COVID-19 merely based on clinical symptoms ^[14]^. We found that patients’ average age trended older after the outbreak ^[15]^. A reasonable explanation for this was that patients with relatively severe diseases had to seek medical care in hospitals even though they faced a high risk of cross-infection with COVID-19. Older people generally have a higher risk of severe disease, but this difference did not exist in critically ill patients, likely because most critical patients were elderly people.

All patients in the FC were regarded as potential sources of infection, thus decreasing the number of patients who had to be transferred to the ED from the FC was an important strategy. Before the outbreak, the major work of the FC was identifying influenza, which might take about half an hour. Once negative results were reported, patients could be transferred to the ED with limited precautions. Once COVID-19 hit, frequent transfers between the FC and ED might cause crowded situations in the ED and increase exposure risks. Our data showed a lower transfer rate after the outbreak, likely due to increasing amounts of medical treatment (as opposed to just testing before the outbreak) in the FC. To those patients who were finally transferred to the ED within 24 hours, a longer FC retention time was observed due to the prolonged screening time for COVID-19. During their time in the FC, patients received treatments aimed at decreasing the number of patients in the ED. Even though some patients had to seek further medical advice in the ED, initial treatments given in the FC might also shorten their length of stay in the ED.

Intensive screening played an important role in COVID-19 identification. Before the outbreak, fever was the only screening indicator. Unfortunately, 11.5% of COVID-19 patients did not manifest fever, but as many as 82.4% of patients had respiratory symptoms, such as cough, expectoration and dyspnea^[12]^. These phenomena impelled us to expand screening criteria. Thus, all patients meeting one or more of the following conditions had to be screened in the FC: positive COVID-19 contact exposure, fever or respiratory symptoms. With this new criteria, the number of FC patients grew dramatically in the 40 days after January 20, 2020.

Multiple testing methods were trialed to decrease false negatives. As COVID-19 has diverse manifestations, it was unreliable to identify this disease based on only one method. In our screening process, multiple methods, including blood cell analysis, chest CT^[16]^, SARS-COV-2 nucleic acid^[17]^ and antibody tests^[18]^ were used to screen for this disease. However, each method had its own false negative phenomena, therefore all patients suspected of any viral infection were recommended to receive another nucleic acid test once more in 24 hours, which helped guarantee that patients who didn’t retest again and were transferred to the ED within 24 hours had an extremely low risk of COVID-19. Additionally, in order to avoid false negative results, patients with the following conditions were suggested to test for SARS-COV-2 nucleic acid once again one week later, even though previous RNA tests were negative twice: (1) confirmed COVID-19 contact history; (2) clinical manifestations and lab tests suggesting a viral infection unexplained by another disease; (3) chest CT strongly indicating a viral pneumonia. With these strict screening criteria, no COVID-19 cases were diagnosed in ED patients previously seen in the FC.

An effective triage strategy also lowered cross-infection risks. As the number of FC patients grew rapidly, cross-infection prevention was a central concern. According to their COVID-19 contact history and clinical severity, patients were allocated to one of three specialized regions of the FC. First, patients with positive contact histories were suggested to keep a person-to-person distance of at least two meters before further history details were investigated, and only when all tests were reported negative could they leave the FC. Second, critical patents identified at the triage counter were immediately admitted to rescue rooms where experienced physicians would provide further assessment and initial resuscitation. Thus, before the results of the screening tests came out,patients were all treated as potentially infected. Once negative results were reported, they could be transferred to the ED resuscitation rooms; otherwise they would continue being treated in the FC or transferred to dedicated receiving hospitals for COVID-19. Third, FC patients could now begin to receive initial treatment(s) as soon as possible without waiting for transfer to the ED.

This study did show that the seven day mortality rate for critically ill patients was higher after the outbreak than before, even though there were no significant differences of the initial treatment time.Although this may be due to the presence of COVID-19 in the patient population after the outbreak, this may also indicate that the longer length of stay in the FC and resuscitation room in the ED after the outbreak may themselves be factors leading to poor outcomes.

Since the outbreak, nineteen FC patients have been confirmed to be positive for COVID-19. It should be noted that all of these positive cases were identified in the FC and received initial treatment there. More importantly, all patients and medical staff in contact with these patients were strictly followed-up for 14 days and no cross-infections were found.

This retrospective study showed the effect of the changes enacted in the FC at the time of the COVID-19 outbreak. The modifications taken in the FC to change the triage, testing, and treatment pathways had a dramatic effect on the FC,as well as offload ED resources.. Although further studies are needed to determine the exact effects of the FC, the lack of cross-contamination events in the ED seem to suggest a possible avenue to EDs around the world to both safeguard their existing ED patients while appropriately caring for potential COVID-19 patients.

## Data Availability

The data information is available at the hospital information system of PUMCH. All of the data used in this study was anonymised before its use.

https://www.who.int/docs/default-source/coronaviruse/situation-reports/20200328-sitrep-68-covid-19.pdf?sfvrsn=384bc74c_2

http://www.nhc.gov.cn/yzygj/s7659/202001/b91fdab7c304431eb082d67847d27e14.shtml

http://www.nhc.gov.cn/yzygj/s7653p/202003/46c9294a7dfe4cef80dc7f5912eb1989/files/ce3e6945832a438eaae415350a8ce964.pdf

## Declarations

### Ethics approval and consent to participate

This retrospective study was approved by The Ethics Committee of the Peking Union Medical College Hospital, the committee’s reference number:S-K1091.

All of the data used in this study was anonymised before its use.

The data information is available at the hospital information system of PUMCH.

Written informed consent was obtained from all participants

### Conflicts of interest

We declare no conflicts of interest.

### Author contributions

JSW and JHL were responsible for the conception and design of the study.JSW,HS, PXS, SYX,YL,JX,and CTW collected the data, LZ and FL were in charge of statistical analysis.LZ, JSW and JHZ took part in drafting the manuscript. JHW,YLand HDZ revised and approved the final version of the manuscript. All authors read and approved the final submitted version.

### Funding

This research received no specific grant from any funding agency in the public, commercial or not-for-profit sectors.

## Acknowledgements

We appreciate each doctor working in the fever clinic that help control the COVID-19. Special thanks to Jinghong Zhang and Joseph Harold Walline for their advice on reviewing manuscript.

## Abbreviations

COVID-19: Corona Virus Disease 2019
SARS-COV-2: Severe Acute Respiratory Syndrome Coronavirus 2
PUMCH: Peking Union Medical College Hospital
ED: Emergency Department
FC: Fever Clinic
CT: Computed Tomography
APACHE II: Acute Physiology and Chronic Health Examination (APACHE) II score
IDR: Infectious Disease Report
NA: Nucleic Acid
SLE: Systemic Lupus Erythematosus

## Notes

### Competing Interest Statement

The authors have declared no competing interest.

### Clinical Trial

no

